# Clinical and Imaging Findings in COVID-19 Patients Complicated by Pulmonary Embolism

**DOI:** 10.1101/2020.04.20.20064105

**Authors:** Ting Li, Gregory Kicska, Paul E Kinahan, Chengcheng Zhu, Murat Alp Oztek, Wei Wu

## Abstract

**Objective:** To describe clinical, and imaging findings including the evolution pattern in COVID-19 pneumonia complicated by pulmonary embolism (PE).

**Methods:** Eleven of 1453 patients with a probable diagnosis of COVID-19 pneumonia were retrospectively selected for the presence of PE. Clinical and laboratory data were recorded. All cross-sectional CT imaging was qualitatively scored for the first 28 days after onset of symptoms.

**Results:** Of 24 patients underwent CTA-PE, 11 were confirmed with PE. All 11 patients developed acute respiratory distress syndrome (ARDS). The pulmonary emboli were most common in segmental and subsegmental pulmonary arteries. We observed an evolution pattern of predominant findings with ground-glass opacities (GGO) to GGO with crazy paving in 3 patients, then to consolidation with linear densities, or to reticulation in 9 patients. Lung cysts or traction bronchiectasis could be seen from day 5 to 9 after symptoms and reticulation, subpleural curvilinear lines were more common from day 20. The pulmonary opacities were predominantly peripheral in distribution with relative sparing of nondependent lungs. The severity of lung involvement was high with an average score of 9.7 in the first phase, 18 in the second phase plateauing in the next two phases, with a slight decrease to 16.9 in the late phase.

**Conclusion:** The incidence of PE among suspected patients in COVID-19 was high. The pulmonary emboli were most common in segmental and subsegmental pulmonary arteries. Our study suggests PE may occur with increased frequency in the ARDS subgroup. The evolution of radiographic abnormalities showed a general pattern, but are also unique with more extensive lung injury and specific imaging features, which may due to the exist of ARDS in these patients.

## Introduction

The outbreak of the novel coronavirus disease (COVID-19) started in Wuhan, China in late December 2019 and rapidly spread to the global world, including more outbreaks in European region and region of America. In the United States, the community transmission of COVID-19 was first detected in February 2020, by now, it has spread to nearly 800,000 people in the U.S. across all 50 states [1]. Computed tomography (CT) has played an important role in the diagnosis of COVID- 19 since the disease outbreak and has also helped identify patients with severe complications. One such complication, pulmonary embolism (PE), has been described as occurring with increased frequency in the setting of influenza infection [2,3]. However, to our knowledge, there is no detailed study describing PE or the clinical and imaging features that may be common to this subgroup of patients in the setting of COVID-19 infection. In this study we describe the clinical presentation, laboratory results and evolution of imaging findings in COVID-19 patients who develop PE. We describe the findings in individual patients and report common trends in the imaging.

## Materials and methods

The Ethics Committee of Wuhan Central Hospital, Tongji Medical College, Huazhong University of Science and Technology approved this retrospective study and informed consent was waived.

### Patients

Eleven patients with documented pulmonary embolism were retrospectively identified from 1453 patients suspected to have COVID-19 between Jan 1 to Feb 13, 2020. All 1453 patients originally presented to our hospital with severe acute respiratory symptoms and the chest CT imaging abnormalities were compatible with viral pneumonia. 24 out of 1453 patients with elevated D-dimer and suspected to have pulmonary embolism were evaluated with CTA-PE. Patients were included in the study if a PE was documented on pulmonary CT angiography (CTA-PE) following diagnosis of confirmed or probable COVID-19 based on WHO guidelines of global surveillance for COVID-19 [4].

### Clinical parameters and laboratory tests

Clinical characteristics including basic demographics, presenting symptoms, past medical history, clinical outcomes were obtained through the electronic medical record. The time interval between onset of symptoms and CT examinations during the first month after symptoms were obtained. Hematological and biochemical blood test results were gathered via the electronic patient record system.

### CT protocol

All the non-contrast CT examinations were performed (Bright Speed 16 CT, GE Healthcare, Japan) under the CT protocol as follows: 120-kVp tube potential; automatic tube current (180 mA-300 mA); iterative reconstruction technique; 20 mm detector, 0.6 second rotation time; section thickness, 5 mm; collimation, 1.25mm; pitch, 1.375; matrix, 512*512; and breath hold at full inspiration. Reconstruction kernel used was lung smooth with a thickness of l.25mm and a slice interval of 1.25 mm.

CTA-PE followed the standard protocol established at our institution for the diagnosis of PE with a section scanner (Brilliance 128ict Philips Medical Systems, U.S.A). After injection of 100ml of nonionic contrast material (300 mg iodine/ml) with a flow of 4.0 ml/sec, scanning was started with a delay of 15 seconds. Typical exposure parameters were 320 mA at 120 kVp with a gantry speed of 0.35 second per rotation. Scanning was performed at full inspiration with a table feed of 5 mm, a slice thickness of 5 mm, and a reconstruction index of 3 mm.

### Image review and interpretation

All available CT images (non-contrast CT and CTA-PE) of the patients were obtained and reviewed by two thoracic radiologists (T.L. and W.W.). Images were evaluated independently and discrepancies were addressed by consensus method. All the CT scans were assessed for the presence and distribution of pulmonary abnormalities. Pulmonary abnormalities were defined according to the accepted glossary of terms in literature [5, 6, 7]. Following findings were recorded: 1) Pulmonary opacities (including the predominant finding, the presence of ground-glass opacities, consolidation, crazy paving, CT halo sign, reverse halo sign, lung cyst(s), air bronchogram, reticulation, linear densities, subpleural parenchymal bands, subpleural curvilinear lines, mosaic attenuation. 2) Airways: bronchial wall thickening, bronchiectasis (excluding traction), traction bronchiectasis. 3) Other findings: nodules, pleural effusion, pericardial effusion, lymphadenopathy. 4) Underlying lung disease: emphysema, fibrosis. 5) Axial distribution: diffuse, peribronchovascular, peripheral distribution, anterio-posterior density gradient. The extent of involvement in each lung was assessed visually using the qualitative scoring system [8] from 0 to 5: 0, no involvement; 1, < 5% involvement; 2, 6%-25% involvement; 3, 26%-49% involvement; 4, 50%-75% involvement; 5, >75% involvement. The total lung involvement score was the sum of total 5 lobes ranging from 0 to 25. The location of the pulmonary emboli was also recorded.

### Statistical Analysis

Quantitative data were presented as median (inter-quartile range, IQR) or mean (range) and the counting data were presented as the percentage of the total numbers in the group. Descriptive analysis was performed with MATLAB (R2014b, MathWorks, Natrick, MA)

## Results

### Patient characteristics and laboratory findings

The demographic and clinical characteristics are summarized in Table 1. 24 patients were evaluated for pulmonary embolism with CTA-PE. 11 of 24 patients had documented pulmonary embolism and were included in the study (7 male and 4 female), with average age of 63 ranging from 36 to 76 years. All the patients were current residents of Wuhan city. Seven patients had history of hypertension, and none had any known underlying pulmonary disease. The most common symptoms were dyspnea (10/11, 90.9%), fever (8/11, 72.7%), chills (7/11, 63.6%), cough (7/11, 63.6%), loss of appetite (6/11, 54.6%), other symptoms including expectoration (4/11, 36.4%), abdominal pain (3/11, 27.3%), myalgia (3/11, 27.3%), throat pain (1/11, 9.1%). Three out of 11 patients died during the hospitalization, all other patients were discharged after treatment.

**Table 1.** Patient clinical characteristics.

| **Clinical characteristics** | **All patients (N=11)** |
| --- | --- |
| **Age** | 63 (36-76) # |
| **Gender** |  |
| Male | 7(63.6%) |
| **Past medical history** |  |
| Hypertension | 7 (63.6%) |
| Schizophrenia | 1 (9.1%) |
| None | 3 (27.3%) |
| **Presenting symptoms** |  |
| Throat pain | 1 (9.1%) |
| Dyspnea | 10 (90.9%) |
| Cough | 7 (63.6%) |
| Expectoration | 4 (36.4%) |
| Chills | 7 (63.6%) |
| Fatigue | 8 (72.7%) |
| Abdominal pain | 3 (27.3%) |
| Loss of appetite | 6 (54.6%) |
| Myalgia | 3 (27.3%) |
| Fever | 8 (72.7%) |
| **Clinical outcome** |  |
| Deceased | 3 (27.3%) |
| Discharge | 8 (72.7%) |

Note: #, data was presented as mean (minimum-maximum); all other counting data were presented as count (percentage of the total)

The blood tests of the patients showed lymphopenia with relatively decreased lymphocyte count (median, 0.84*109cells/Liter) and a lower lymphocyte percentage (median, 8%). The biochemical blood tests showed elevated C-reactive protein lever (median, 6.86mg/dL) in 9 patients and D-Dimer (median, 26.91µg/ml) in all patients. All the patients were diagnosed with acute respiratory distress syndrome (ARDS) according to WHO interim guidance for severe acute respiratory infection of COVID- 19[9].

Of the 11 patients that were clinically diagnosed to have COVID-19 infection, only four had positive reverse-transcription polymerase chain reaction (RT-PCR) test. In individuals with a negative RT-PCR test, a presumptive diagnosis of COVID-19 infection was made in accordance with the Chinese Health Commission case definition for COVID-19 (fifth edition) which used the high pretest probability conferred by the strong epidemiological link to Wuhan, unexplained acute respiratory symptoms, and typical imaging, and absence of evidence of other respiratory pathogens [10].

### Imaging interpretation

After evaluation, the CT scans were categorized according to the time from onset of the symptoms as phase 1 to phase 4 with interval of 5 days in the first 20 days, and phase 5 was defined from Day 20 to 28. The timeline of the CT scans in these patients was showed in Figure 1. In the first 15 days (phase 1 to phase 3), the most common CT findings were GGO with or without crazy paving pattern in the first 15 days. In the second half of the month, the most common predominant CT findings were consolidation, reticulation and linear densities. In 3 out of 11 patients, the pulmonary abnormalities started with GGO, progressed to crazy paving pattern with superimposed inter- and intralobular septal thickening, in 9 out of 11 patients, the GGO with crazy paving pattern evolved to consolidation with linear densities or reticulation. The predominant findings in different phases for each patient was presented in Table 2. Of the 11 patients, 10 (90.91%) had GGO with crazy paving sign, which was more frequent in the first 15 days while 8 (72.7) had consolidations or linear densities, which was more frequent in the second half of the month. Four patients (36.4%) had reticulation which was found mostly in the last phase, six patients (54.6%) had subpleural curvilinear lines which was also found mostly in the last phase. Three patients had lung cysts in day 7 to day 19 and traction bronchiectasis was also found in 2 patients in day 10 to day 28. CT halo sign and reverse halo sign were absent in all the patients. Six patients (54.6%) had pleural effusions in this study. None of the patients had pulmonary nodules, lymphadenopathy or other preexisting lung disease. The peripheral distribution of the opacities was most common (9/11, 81.8%), and the anterior-posterior density gradient was found in 7 patients (63.6%). All the lobes in the lung were involved in these patients. The mean total involvement score was 9.7 (SD=0.7) in phase 1, 18.2 (SD=0.6) in phase 2, 18.1 (SD=0.4) in phase 3, 18.3 (SD=0.3) in phase 4 and 16.9 (SD=0.3) in phase 5. The details of the CT findings in each phase was summarized in Table 3. The total lung involvement score for each patient and the average score during the first 28 days were shown in Fig. 2. There were 3 patients with PE in lobar, segmental and subsegmental pulmonary arteries, 4 patients with PE in segmental and subsegmental pulmonary arteries and 4 patients with PE in subsegmental pulmonary arteries only. The location of the PE for each patient is shown in Table 4. The lower-limb ultrasonography was performed in 2 out of 11 patients and no lower extremity deep vein thrombosis was found.

**Table 2.**
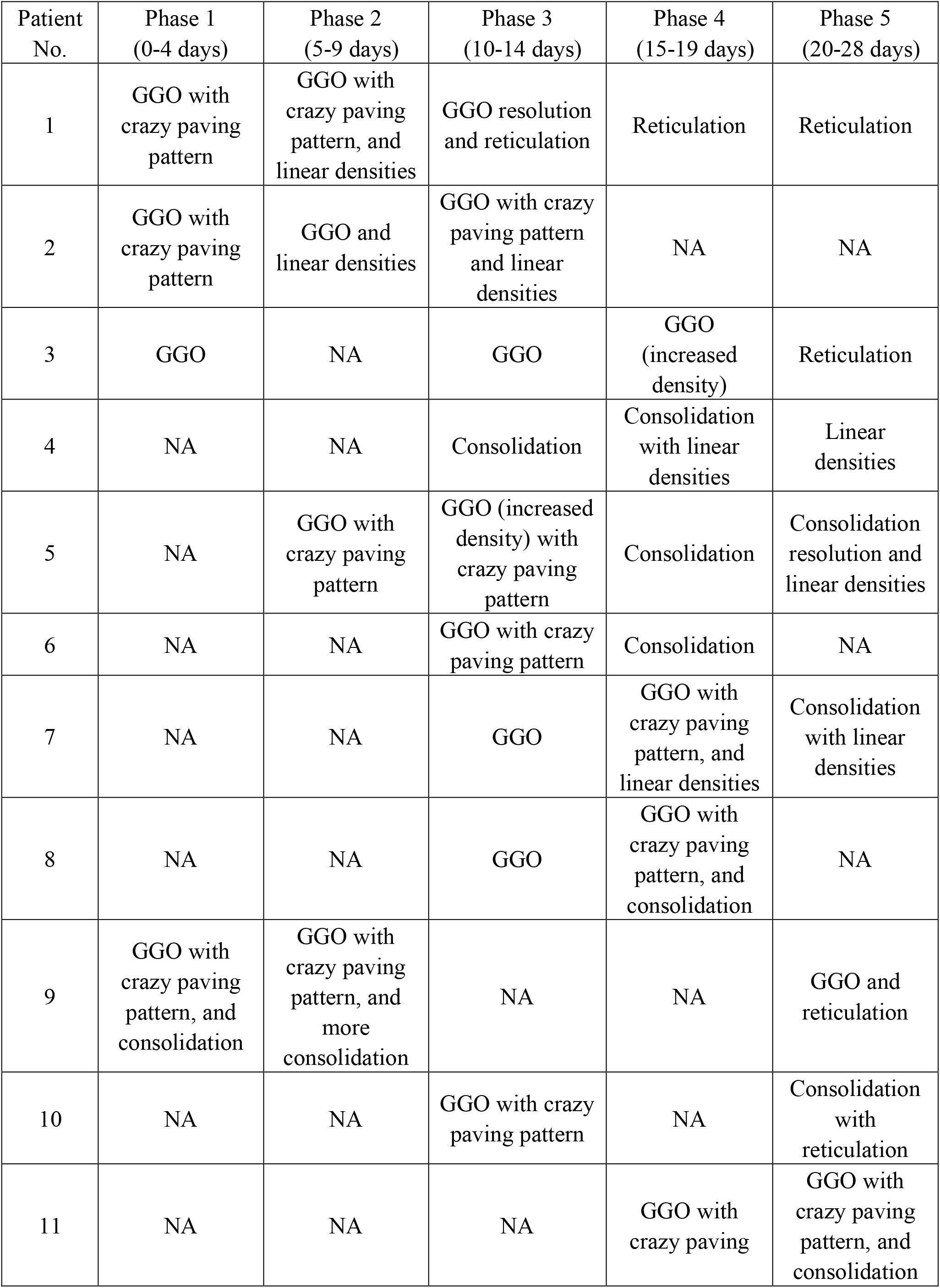
Predominant longitudinal CT findings in 11 patients during the 1^st^ month.

| Patient No. | Phase 1 (0-4 days) | Phase 2 (5-9 days) | Phase 3 (10-14 days) | Phase 4 (15-19 days) | Phase 5 (20-28 days) |
| --- | --- | --- | --- | --- | --- |
| 1 | GGO with crazy paving pattern | GGO with crazy paving pattern, and linear densities | GGO resolution and reticulation | Reticulation | Reticulation |
| 2 | GGO with crazy paving pattern | GGO and linear densities | GGO with crazy paving pattern and linear densities | NA | NA |
| 3 | GGO | NA | GGO | GGO (increased density) | Reticulation |
| 4 | NA | NA | Consolidation | Consolidation with linear densities | Linear densities |
| 5 | NA | GGO with crazy paving pattern | GGO (increased density) with crazy paving pattern | Consolidation | Consolidation resolution and linear densities |
| 6 | NA | NA | GGO with crazy paving pattern | Consolidation | NA |
| 7 | NA | NA | GGO | GGO with crazy paving pattern, and linear densities | Consolidation with linear densities |
| 8 | NA | NA | GGO | GGO with crazy paving pattern, and consolidation | NA |
| 9 | GGO with crazy paving pattern, and consolidation | GGO with crazy paving pattern, and more consolidation | NA | NA | GGO and reticulation |
| 10 | NA | NA | GGO with crazy paving pattern | NA | Consolidation with reticulation |
| 11 | NA | NA | NA | GGO with crazy paving | GGO with crazy paving pattern, and consolidation |

**Table 3.** CT findings and imaging signs of pulmonary opacities during the 1^st^ month.

|  | Total patients (N=11) | 0-4 days (N=3) | 5-9 days (N=5) | 10-14 days (N=9) | 15-19 days (N=7) | 20-28 days (N=8) |
| --- | --- | --- | --- | --- | --- | --- |
| Pulmonary opacities |  |  |  |  |  |  |
| GGO | 10 (90.9) | 3 (100) | 5 (100) | 9 (100) | 4 (57.1) | 4 (50) |
| Consolidation | 8 (72.7) | 0 | 2 (40) | 4 (44.4) | 3 (42.9) | 5 (62.5) |
| Crazy paving sign | 10 (90.9) | 2 (66.7) | 3 (60) | 5 (55.6) | 3 (42.9) | 1 (12.5) |
| Halo sign | 1 (9.1) | 0 (0) | 0 (0) | 0 (0) | 0 (0) | 0 (0) |
| Reverse halo sign | 0 (0) | 0 (0) | 0 (0) | 0 (0) | 0 (0) | 0 (0) |
| Lung cyst | 3 (27.3) | 0 (0) | 1 (20) | 3 (33.3) | 2 (28.6) | 0 (0) |
| Air bronchogram | 5 (45.5) | 2 (66.7) | 1 (20) | 2 (18.2) | 2 (28.6) | 0 (0) |
| Reticulation | 4 (36.4) | 0 (0) | 1 (20) | 1 (11.1) | 1 (14.3) | 4 (50) |
| Linear densities | 8 (72.7) | 0 (0) | 2 (40) | 3 (33.3) | 4 (57.1) | 7 (87.5) |
| Subpleural parenchymal bands | 2 (18.2) | 1 (33.3) | 1 (20) | 0 (0) | 1 (14.3) | 2 (25) |
| Subpleural curvilinear lines | 6 (54.6) | 1 (33.3) | 1 (20) | 1 (11.1) | 3 (42.9) | 6 (75) |
| Mosaic attenuation | 1 (9.1) | 0 (0) | 0 (0) | 0 (0) | 1 (14.3) | 1 (12.5) |
| Airways |  |  |  |  |  |  |
| Bronchial wall thickening | 0 | 0 (0) | 0 (0) | 0 (0) | 0 (0) | 0 (0) |
| Bronchiectasis | 0 | 0 (0) | 0 (0) | 0 (0) | 0 (0) | 0 (0) |
| Traction bronchiectasis | 2 (18.2) | 0 (0) | 0 (0) | 1 (11.1) | 0 (0) | 1 (12.5) |
| Other findings |  |  |  |  |  |  |
| Nodules | 0 (0) | 0 (0) | 0 (0) | 0 (0) | 0 (0) | 0 (0) |
| Pleural effusion | 6 (54.6) | 0 (0) | 0 (0) | 5 (55.6) | 3 (42.9) | 4 (50) |
| Pericardial effusion | 1 (9.1) | 0 (0) | 0 (0) | 1 (11.1) | 0 (0) | 1 (12.5) |
| Lymphadenopathy | 0 | 0 (0) | 0 (0) | 0 (0) | 0 (0) | 0 (0) |
| Underlying lung disease |  |  |  |  |  |  |
| Emphysema | 0 (0) | 0 (0) | 0 (0) | 0 (0) | 0 (0) | 0 (0) |
| Fibrosis | 0 (0) | 0 (0) | 0 (0) | 0 (0) | 0 (0) | 0 (0) |
| Axial distribution |  |  |  |  |  |  |
| Diffused | 3 (27.3) | 0 (0) | 1 (20) | 2 (18.2) | 3 (42.9) | 1 (12.5) |
| Peribronchovascular distribution | 4 (36.4) | 2 (66.7) | 1 (20) | 2 (18.2) | 1 (14.3) | 1 (12.5) |
| Peripheral distribution | 9 (81.8) | 3 (100) | 4 (80) | 8 (88.9) | 4 (57.1) | 6 (75) |
| Anterio-posterior density gradient | 7 (63.6) | 2 (66.7) | 2 (40) | 5 (55.6) | 1 (14.3) | 0 (0) |
| Lobe involvement score |  |  |  |  |  |  |
| LUL | NA | 1.3 | 3.2 | 3.3 | 3.3 | 3.1 |
| LLL | NA | 2.3 | 4.2 | 3.9 | 3.9 | 3.5 |
| RUL | NA | 2.3 | 3.6 | 3.5 | 3.9 | 3.5 |
| RML | NA | 1 | 3 | 3.2 | 3.3 | 3.0 |
| RLL | NA | 2.7 | 4.2 | 4.2 | 4.0 | 3.8 |
| total | NA | 9.7 | 18.2 | 18.1 | 18.3 | 16.9 |

Note: data was presented as count (percentage of the total)

**Table 4.** Location of Pulmonary embolism in all 11 patients.

| Patient No. | PE Location |
| --- | --- |
| 1 | Lobar, segmental and subsegmental LUL; lobar, segmental and subsegmental LLiL; lobar, segmental and subsegmental LLL; lobar, segmental and subsegmental RUL; lobar, segmental and subsegmental RML; lobar, segmental and subsegmental RLL |
| 2 | Segmental LLL; subsegmental LUL; subsegmental RUL; subsegmental RLL |
| 3 | Segmental and subsegmental RLL |
| 4 | Lobar, segmental and subsegmental LLL; segmental LUL; subsegmental LLiL; segmental and subsegmental RLL |
| 5 | Segmental and subsegmental RML; Segmental and subsegmental RLL |
| 6 | Subsegmental LUL |
| 7 | Lobar, segmental and subsegmental LLL; segmental and subsegmental RLL; subsegmental RML |
| 8 | Subsegmental RUL; subsegmental LUL |
| 9 | Subsegmental LLL |
| 10 | Segmental and subsegmental LUL; subsegmental LLL; segmental and subsegmental RLL |
| 11 | Subsegmental RLL |

**Figure 1.**
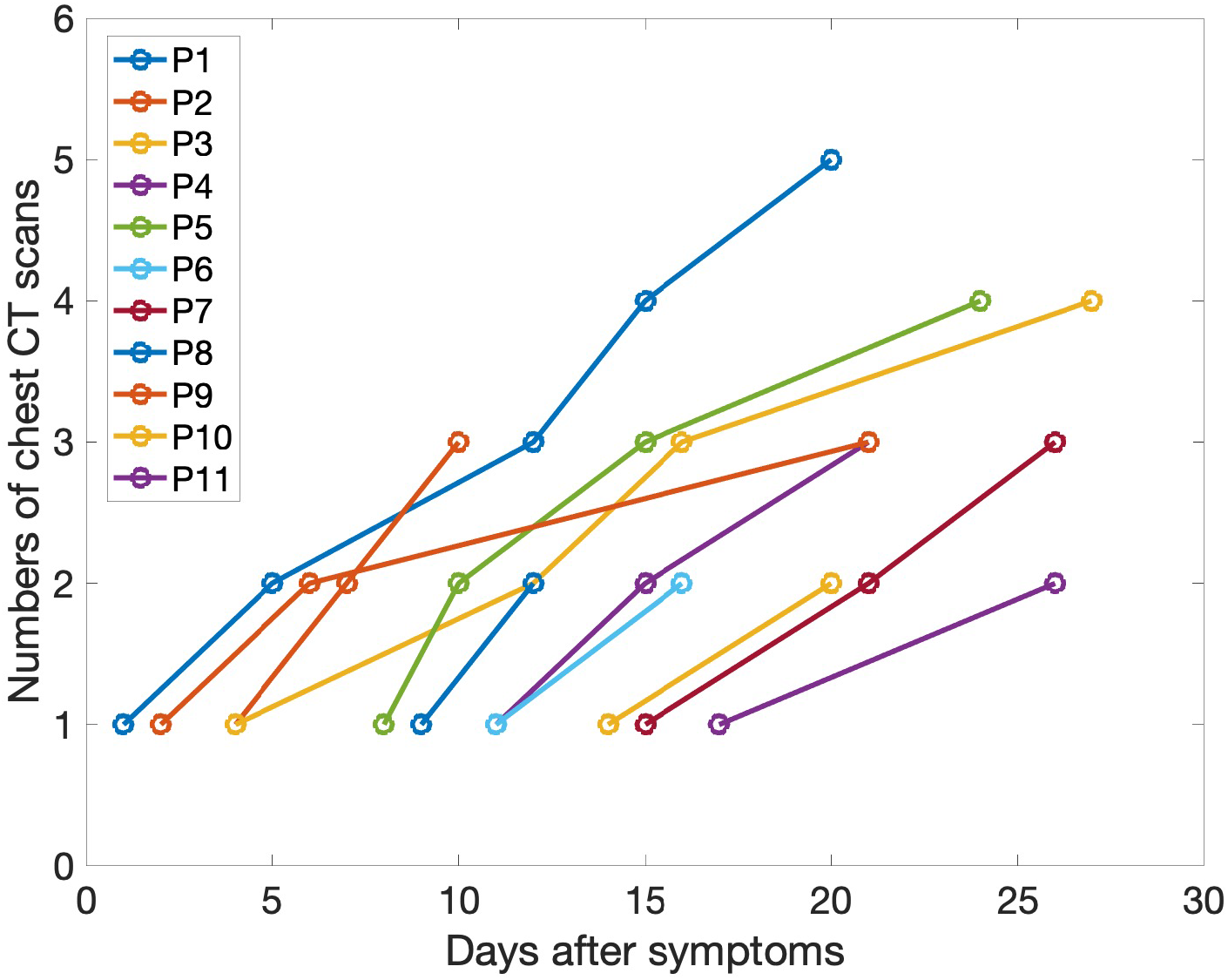
Timeline of the chest CT scans in all the patients during the first 28 days after onset of symptoms.

**Figure 2.**
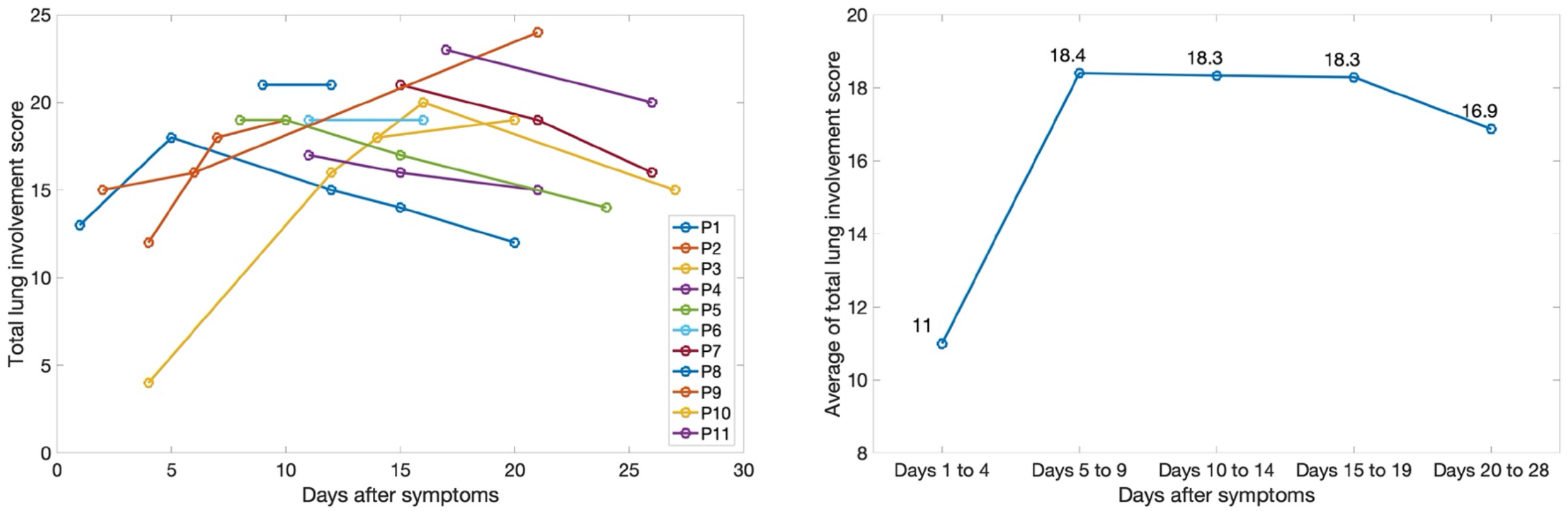
Longitudinal total lung involvement score for each patient (left side) and the average score (right side) during the first 28 days.

## Discussion

5% of patients with COVID-19 infections develop severe complications and become critically ill [11], however, to our knowledge, there was just two case reports about the complication of acute pulmonary embolism in two COVID-19 patients [12,13]. It is important to report the clinical and imaging features of this patient’s subgroup, because it may suggest a unique pattern that predicts PE or other complications. In this case series, we described the clinical findings as well as the evolution of imaging features in 11 COVID-19 patients who developed pulmonary embolism.

Unlike the previous reported studies of fever being the most common symptoms, the most common onset symptoms for patients in our study was dyspnea. All the patients presented lymphopenia in hematological blood test indicating viral infection. Nine of the 11 patients had a higher C-reactive protein and all the patients had increased blood level of D-dimer (26.91µg/ml (IQR 18.02-43.84)), which was higher than in other reported studies [15,16]. We observed an evolution pattern of predominant findings with GGO to GGO with superimposed inter- and intralobular septal thickening in 3 patients (crazy paving pattern), then to consolidation with linear densities, or to reticulation in 9 patients (Figure. 3). Lung cysts, traction bronchiectasis could be seen from day 7 from onset of symptoms and reticulation, subpleural curvilinear lines were more common from day 20 (Figure 4 and Figure.5). In addition to the peripheral lung distribution of the pulmonary opacities, which was similar to a recently published study [14], in our cohort anterior-posterior density gradient and relative sparing of the non-dependent lung were also seen in the first 15 days (Figure 4). It has been reported that the anterior-posterior density gradient is typical in the early phase of ARDS, due to the posterior compressive atelectasis caused by the weight of the overlying parenchyma [15, 16]. The lung cysts were reported to be features of the later stages of ARDS, which may be caused by prolonged ventilation or secondary to pneumonia.

**Figure 3.**
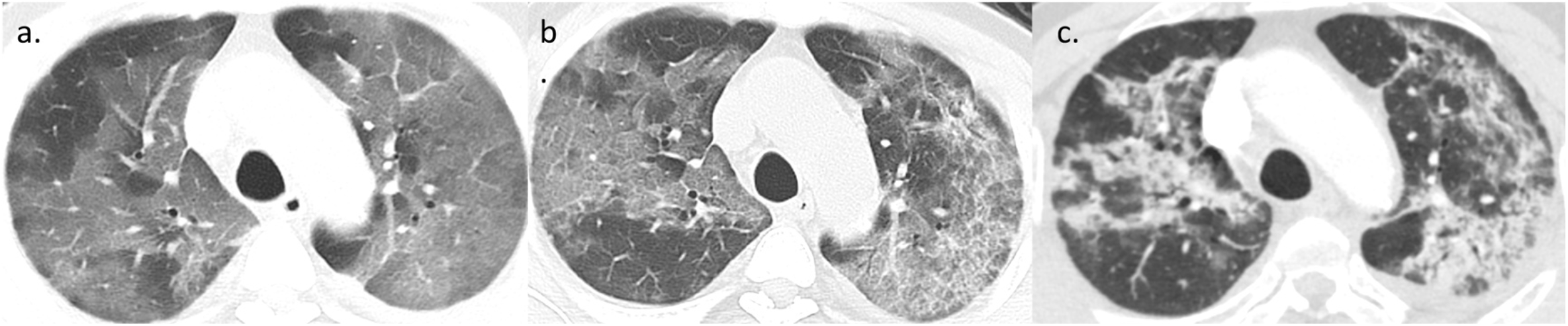
A patient with confirmed diagnosis of COVID-19. a. The images at day 15 from onset of symptoms showed the predominant ground-glass opacities in bilateral lungs. b. In day 21, the ground-glass opacities were superimposed with inter- and intralobular septal thickening, presenting a crazy paving pattern. c. In day 26, the previous ground-glass opacities evolved to consolidation with some linear densities.

**Figure 4.**
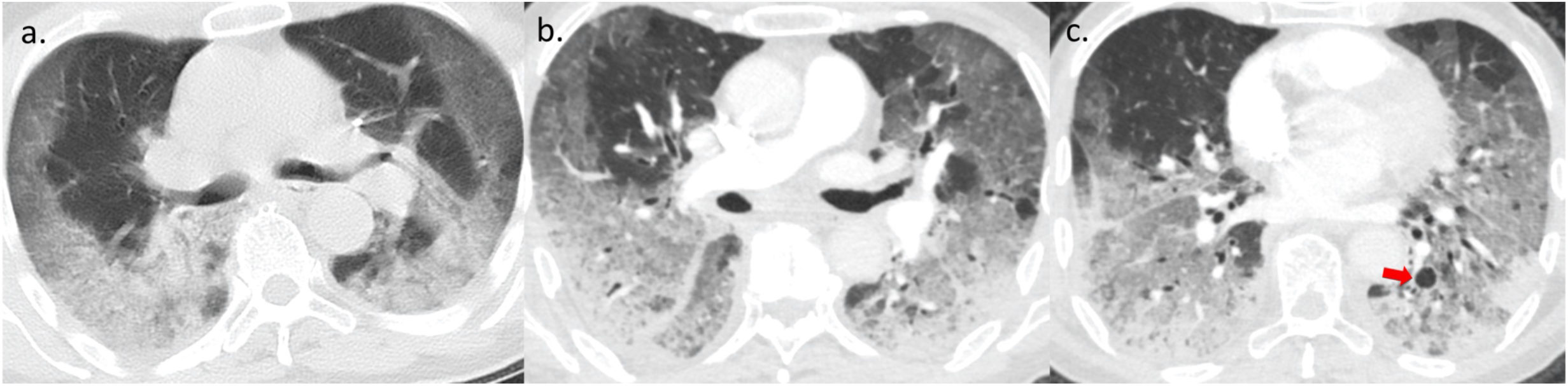
A patient with probable diagnosis of COVID-19. a. The images at day 9 from onset of symptoms showed the predominant ground-glass opacities in peripheral distribution with the anterior-posterior density gradient. b. In day 12, the ground-glass opacities were superimposed with inter- and intralobular septal thickening, and with more dense consolidations in the dependent regions widespread ground-glass opacities and relatively normal parenchyma in non-dependent areas. c. In day 12, A lung cyst in left lower lobe was seen.

**Figure 5.**
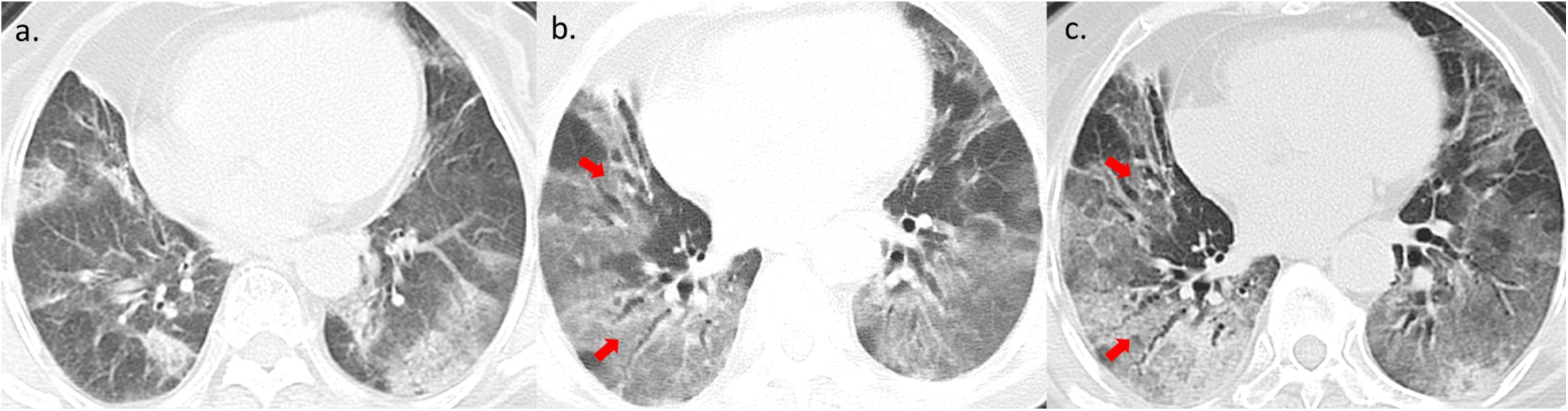
A patient with probable diagnosis of COVID-19. a. The images at day 4 from onset of symptoms peripheral scattered ground-glass opacities. b. In day 7, the ground-glass opacities were more widespread with traction bronchiectasis was seen in right lower lobe (arrow). c. In day 10, traction bronchiectasis (arrow) was more apparent, the anterior-posterior density gradient was also seen.

Unlike those patients in previous imaging studies [14,17], more than half of the patients in our study had pleural effusion. Compared to the lung involvement score in another study using the same qualitative scale of COVID-19 [18] with the average score range 2 to 7, the severity of lung involvement in our study was high with an average score of 9.7 in the first phase, 18 in the second phase plateauing in the next two phases, with a slight decrease in the average score to 16.9 in the late phase indicating a little bit of resolution. It is evident that the severe lung injury started early and persisted in extent and severity beyond 20 days from the onset of symptoms.

It’s reported that the proportion of cases diagnosed with PE among suspected patients has steadily decreased to 5-20% due to the use of D-dimer measurement [19]. In our study, pulmonary embolism was documented in 11 (46%) of 24 patients with higher D-dimer and suspected of PE, which is much higher than expected frequency for this patient population. Two out of 11 patients had lower-limb ultrasonography performed and no lower extremity deep vein thrombosis was found. Acute infection was reported to be a strong trigger for venous thromboembolism independent of concomitant immobilization [20], and there was higher risk estimates for infection preceding pulmonary embolism (PE) than deep vein thrombosis, and respiratory tract infection had a greater impact on venous thromboembolism than non-respiratory infections [21]. In our study, all the patient developed pulmonary embolism after severe COVID-19 pneumonia without major risk factors for PE including underlying cardiopulmonary diseases, oncologic disease history, past deep vein thrombosis, or prolonged immobility more than 3 days, which may further confirm the role of infection as an independent trigger for thromboembolism. It was also reported that vascular hyaline thrombi was found in alveolar interstitium based on the limited autopsy cases of COVID-19 in China [22]. The pulmonary emboli in our case series were most commonly observed in segmental and subsegmental pulmonary arteries, and there were four patients with PE in subsegmental arteries only. In the segmental and subsegmental pulmonary arteries, the emboli were scattered and sometimes presented as minor filling defects (Figure. 6), which may be caused by the microthrombi formation in the lungs. However, whether local hyper-inflammatory cytokine responses in the pulmonary circulation in COVID-19 lead to the coagulation activation cascade and thrombus formation in situ still remained open, interesting and warranted further investigation.

**Figure 6.**
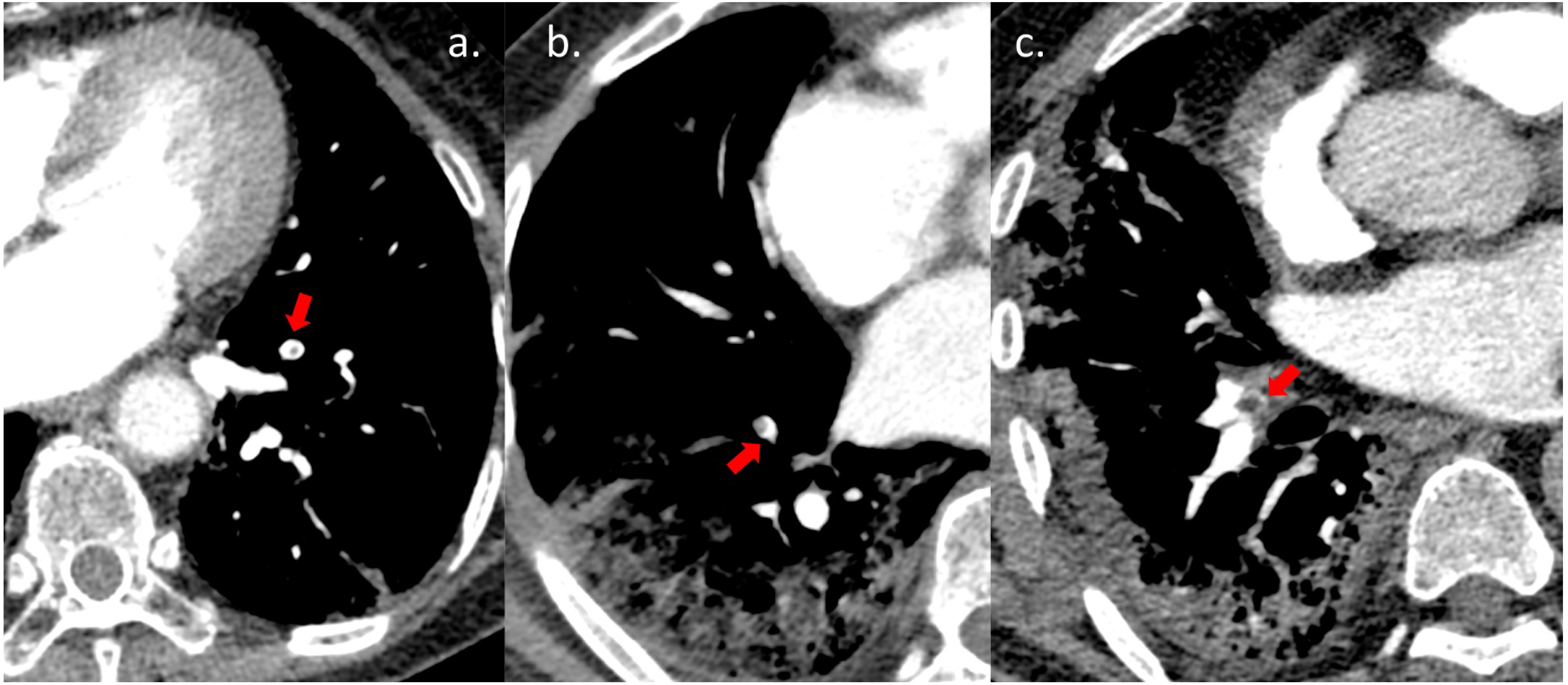
a. A patient with probable diagnosis of COVID-19, a small filling defect was seen in subsegmental LLL artery. b. A patient with probable diagnosis of COVID-19, a filling defect was seen in subsegmental RLL artery c. A patient with confirmed diagnosis of COVID-19, the beginning of the medial basal segmental artery in RLL was blocked.

Development of acute respiratory distress syndrome (ARDS) in COVID-19 patients plays a significant role in mortality. In a recent study of 1099 patients with COVID- 19, 3.4% developed ARDS [11], and in a smaller study of 138 hospitalized patients, 8% developed ARDS [23]. In the critically ill patients, 67% of patients developed ARDS, which was the most common complication. And in critically ill patients with ARDS, 61.5% died within 28 days [24].

All patients in this study developed ARDS, suggesting a correlation between ARDS and PE. This may be explained by the proposed mechanism of thrombo-inflammation causing ARDS, which is inflammation that activates platelets and causes damage to the endothelium, resulting in fibrin deposition and thrombus formation [25]. In a mouse models of ARDS, infection with the influenza virus increases platelet aggregation, pulmonary microvascular thrombosis, endothelial damage and the hyper-inflammatory cytokine responses [26,27,28]. Dysregulation of these same physiologic responses have been associated with pulmonary embolism (PE) [29], and prior studies have described PE in the setting of influenza infection [2,3].

Our study has several limitations: First, only 11 patients were used in this analysis. However, the population from which these patients were selected represents approximately 1.5% of all COVID-19 patients reported worldwide. A larger data set could be achieved with a multicenter study or in the future as more patients present. A second limitation is that the CT scans in different time phases were not equally represented. For example, there were fewer CT scans performed in the first 10 days after onset of symptoms compared other time periods.

A third limitation is that only 4 out of 11 patients were RT PCR test positive. Although a positive RT-PCR test is not required for diagnosis based on current guidelines in Hubei province, the test is considered highly specific, increasing confidence in the diagnosis. COVID-19 virus load is higher in early days after infection [30], and the limited number of the RT-PCR kits available delayed testing for many the patients, causing the test to be performed when it is less accurate. Patient with negative RT-PCR would be retested in 3 days, but 3 of the patients died before this was possible, also suggesting they presented at a later stage of infection. RT-PCR is not required for diagnosis and this is supported by multiple studies. For example, on study showed that 60% to 93% of patients with an initial positive chest CT consistent with COVID-19 had negative RT-PCR results [31]. In another case series, the sensitivity of CT for COVID-19 diagnosis was 98% compared to RT-PCR sensitivity of 71% within 3 days from onset of symptoms [32]. Although 7 of the patients in this study had negative RT-PCR results, they were still probable COVID- 19 according to WHO’s interim guidance and clinically confirmed COVID-19 according to the Chinese Guideline of Diagnosis and Treatment for COVID-19, which was only applicable in Hubei Province due to the insufficient sensitivity of RT-PCR testing in the epidemic center [10].

In conclusion, our case series summarizes the clinical and imaging findings of 11 COVID-19 patients complicated PE. Pulmonary embolism was documented in 11 of 24 patients suspected of PE, higher than expected frequency for this patient population. All patients in our study developed ARDS, suggesting PE may occur with increased frequency in the ARDS subgroup. This high incidence of PE in COVID-19 patients has not been reported previously. The evolution of pulmonary imaging findings shares some common characteristics with non-severe COVID-19 patients, but are also unique in that there is more extensive lung injury, and a longer time for the abnormalities to resolve. Given that all patients with PE developed ARDS, there should be a lower threshold for suspecting PE in COVID-19 patients with ARDS. This may facilitate better detection of PE and reduce mortality and morbidity in COVID-19 patients.

## Data Availability

Data was not available publicly bu can request via contacting with corresponding author.

## Acknowledgement

We thank Sudhakar, Pipavath in University of Washington for his valuable contribution in editing the manuscript.

